# Beneath the surface: mental health, and harassment and abuse of athletes participating in the FINA (Aquatics) World Championships, 2019

**DOI:** 10.1101/2021.04.06.21254987

**Authors:** Margo Mountjoy, Astrid Junge, Christer Magnusson, Farhad Moradi Shahpar, Edgar Ivan Ortiz Lizcano, Josip Varvodic, Xinzhai Wang, Mohamed Yahia Cherif, Lee Hill, Jim Miller

**Author notes:** **Contact for corresponding author:** Dr. Margo Mountjoy, McMaster Medical School, 10-B Victoria Street South, Kitchener, ON, Canada N2G 1C5. **Patient and Public Involvement:** Athlete representatives and members of the FINA Athletes Commission were involved in determining the research priority of mental health and safeguarding, as well as actively recruiting participants and promoting the research project during the FINA World Championships. We will invite athletes and the entourage to help with dissemination and application of the results. **Scientific contributions of each author: Margo Mountjoy, MD, PhD:** Substantial contributions to the conception and design of the study, collection and interpretation of data, drafting, writing, and revising of the manuscript, and approval of the final version to be published. **Astrid Junge, PhD:** Substantial contributions to the conception and design of the study, analysis and interpretation of data, drafting, writing, and revising of the manuscript, and approval of the final version to be published. **Christer Magnusson, MD, Farhad Moradi Shapar, MD, PhD, Edgar Ivan Ortiz Lizcano, MD, Josip Varvodic, MD, Xinzhai Wang, MD, Mohamed Yahia Cherif MD:** Contributions to data collection and approval of the final version to be published. **Lee Hill, PhD:** Substantial contributions to data collection and approval of the final version to be published. **Jim Miller, MD, FAAFP/Sports Medicine:** Substantial contributions to conception of the study, as well as data collection and approval of the final version to be published. All authors agree to be accountable for all aspects of the work in ensuring that questions related to the accuracy or integrity of any part of the work are appropriately investigated and resolved. **Funding:** The data collection was funded by the Federation Internationale de Natation (FINA), Lausanne, Switzerland. FINA had no role in the study design, the collection, analysis, and interpretation of the data, writing the report, or the decision to submit the article for publication. No other funding was received for the present study. **Competing Interest:** None declared. **Ethics approval:** The present study had ethic approval from McMaster University - Hamilton Integrated Research Ethics Board, Canada. **No Additional data:** No supplementary data.

## Abstract

**Objective:** To assess the mental health and experience of sport-related harassment and abuse of athletes participating in the FINA World Championships 2019, and to analyse it in relation to gender and the aquatic disciplines.

**Design:** Cross-sectional study using an anonymous questionnaire.

**Methods:** During the Championships, registered athletes (swimmers, divers, high divers, water polo players, artistic swimmers, open-water swimmers) completed a survey including the main outcome measures of depression (CES-D-10), eating disorders (BEDA-Q), the subjective need for psychotherapeutic support, and the experience of harassment and/or abuse in their sports environment.

**Results:** A quarter of the athletes (n=62, 24.6%) were classified as depressed and more than a third (n=111, 35.0%) as having an eating disorder. More than 40% of the athletes stated that they wanted or needed psychotherapeutic support for mental health problems. Fifty-one athletes (14.9%) had experienced harassment/abuse in sport themselves, and 31 (9%) had witnessed it in another athlete. The experiences of harassment and abuse ranged from unwanted comments about body or appearance (40.2%) to rewards in sport for sexual favours (2.5%) and rape (0.3%). Athletes who had experienced harassment/abuse in sport themselves had higher average scores for depression and eating disorders, and more of them needed psychotherapeutic support. Up to a third would not talk to anybody if they saw or experienced harassment/abuse, and less than 20% would talk to an official for help.

**Conclusion:** Targeted initiatives are required to address the burden of mental health issues and harassment and abuse in sport in the FINA aquatic disciplines.

## INTRODUCTION

> *“I didn’t want to be in the sport anymore* … *I didn’t want to be alive anymore*.” Michael Phelps 2018^1^

The sport community was shocked when Michael Phelps, the most decorated Olympian of all time, first spoke about his struggles with mental health issues in 2016.^2^ A systematic review and meta-analysis of mental health of elite athletes revealed a prevalence of 33.6% for anxiety/depression, 26.4% sleep disturbance, 19.6% symptoms of distress, 9% for alcohol misuse, and 1%–28% for adverse eating habits or eating disorder.^3^ There is a well-established correlation between mental health symptomatology and experience with harassment/abuse both inside and outside of sport.^4^ In athletics, a cross-sectional study of 198 elite Swedish athletes, suicidal ideation of the female athletes was associated with having been sexually abused combined with signs of psychological vulnerability.^5^ A cohort study on Canadian national team athletes demonstrates statistically significant correlations between harassment/abuse in sport and mental health outcomes defined as self-harm, disordered eating and eating disorders, suicidal thoughts and seeking help for mental health issues.^6^

While the original IOC publication on sexual harassment/abuse in sport was published in 2007,^7^ the IOC first implemented safeguarding against harassment/abuse at the 2016 Olympic Games,^8^ and published an updated consensus statement.^9^ A research project was conducted at the Youth Olympic Games 2018 to assess the participating elite youth athletes’ understanding and perception of harassment/abuse in their respective sports. About a third stated that it ‘very likely’ or ‘likely’ occurred in their sport.^10^ This study is the only publication on harassment/abuse in a large cohort of international multi-sport elite athletes.

While there has been a recent increase in scientific publications on mental health and harassment/abuse in sport, there remains a dearth of scientific evidence of the elite aquatic athletes’ experience in these domains. In order to address this gap in knowledge, the objectives of this study were to assess the athletes’ mental health problems and their experience of sport-related harassment/abuse, and to analyse these findings in relation to gender and the aquatic disciplines.

## MATERIALS AND METHODS

An anonymized survey for accredited athletes was performed during the FINA World Championships (WC), 2019 using the same recruitment and data collection procedures as in previous studies.^11-13^

The **questionnaire** started with questions on gender, age, and aquatic discipline, followed by two questions on the extent of health complaints and their effect on performance or participation in the four weeks prior to the WC (based on the Oslo Trauma Research Centre Questionnaire^14^).

**Mental health** was assessed using the 10-item version of the Center for Epidemiological Studies Depression Scale revised (CESD-R-10),^15,16^ the Brief Eating Disorders in Athletes Questionnaire (BEDA-Q)^17^ and the question “Have you ever wanted or needed support from a psychotherapist for personal or mental health problems?” (no/ yes, previously / yes, currently).^18^

The section on **harassment and abuse** was newly developed based on the review of the literature.^10,19^ It consisted of two introductory questions "Do you think harassment and/or abuse occurs in your sport?” (no/not likely/not sure/likely/very likely) and "Have you ever witnessed or experienced yourself any forms of harassment or abuse in your sports environment?” (no/yes, experienced myself/yes, witnessed in another athlete) followed by a table with 13 situations, for each the athlete should report whether or not they had witnessed or experienced it themselves, and if so, who did it (coach/another athlete/others). For details see Table 1.

**Table 1:**
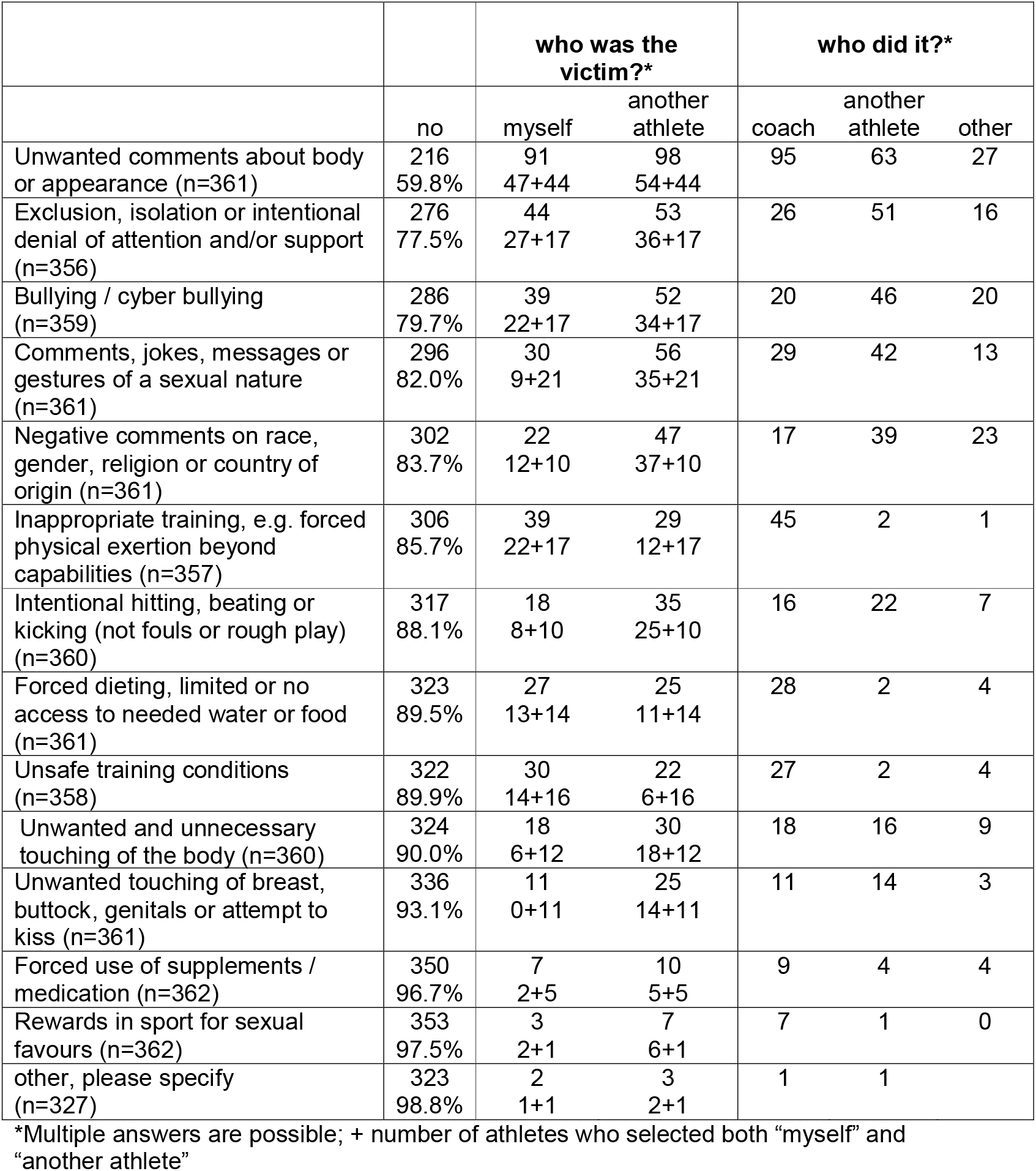
Athlete responses to the question: “Have you ever witnessed or experienced yourself any of the following situations in your sports environment? If yes, please answer both questions.”

The data were processed Excel (Microsoft Office 2011) and SPSS (V.23, IBM). The results are reported using means with standard deviation or frequencies with percentage, and in accordance with STROBE guidelines. We applied ANOVA, t- and Chi^2^-test statistical methods with an accepted significance at p< 0.05.

Support for travel to the FINA WC for data collection was funded by the FINA.

Although FINA approved the project, it had no oversight or influence over the data collection, analysis, interpretation of results or drafting of the manuscript. Ethical design followed the principles of the Declaration of Helsinki for experiments involving humans, including informed consent and observation of privacy rights; ethical approval was granted by the Hamilton Integrated Research Ethics Board.

## RESULTS

A total of 377 athletes participating in the 2019 FINA WC answered the questionnaire (response rate: 14.8%). The 229 (61.1%) female and 146 (38.9%) male athletes were on average 22.4 years old (SD=5.37; range: 14-56), with female participants (21.4, sd=4.71) being significantly younger than male (23.9; sd=5.93; t=4.48, p<.001). The majority of participants were swimmers (SW, n=132, 36.2%), water polo players (WP, n=82, 22.5%) and artistic swimmers (AS, n=79, 21.6%) followed by divers (DV, n=43, 11.8%), high divers (HD, n=21, 5.8%) and open water swimmers (OWS, n=8, 2.2%).

In the 4 weeks prior to the WC, half of the athletes had no symptoms/complaints (n=181), nor any ***injury, illness or other health*** problems that affected their performance or participation (n=187; 51.8%). About a third reported minor health problems (n=125; 34.5%), 40 (11.0%) moderate and 16 (4.4%) severe. More than a quarter of the athletes (n=100; 27.7%) reported minor, 51 (14.2%) moderate and 16 (4.4%) major effects of health problems on their performance; seven (1.9%) could not participate at all.

Based on their answers to the CES-D-10 (n=268, mean=7.38, sd=4.75, range 0-25), a quarter of the athletes (n=62, 24.6%) were classified as ***depressed*** without significant differences between genders. The depression score correlated significantly with the extent of symptoms/health complaints (r=.30, p<.001) and their effect on performance in the four weeks prior to the WC (r=.28, p<.001).

Based on their answers to the six BEDA-Q questions (n=317, mean=0,67, sd=0.60, range 0-3), more than a third of the athletes (n=111, 35.0%) had an ***eating disorder***. The proportion of athletes with an eating disorder differed significantly between females and males (40.2% vs 27.3%; Chi^2^=5.5, p<.05). Athletes with an eating disorder had a significantly higher depression score (F=4.8; p<.05). Thirty athletes (11.6%) were classified to have both depression and eating disorder. About a quarter of the athletes (n=90, 25.1%) stated that they were *trying to lose weight now* (i.e. during the WC), with significantly more females than males (31.7% vs 14.8%; Chi^2^=12.6, p<.001). Sixty athletes (17.4%) had tried to lose weight more than five times.

More than 40% of the athletes stated that they “wanted or needed ***psychotherapeutic support*** for personal or mental health problems” previously (n=103, 28.8%) or currently (n=57,15.9%) without differences across genders or ages. Athletes with and without a subjective need for psychotherapeutic support differed significantly (F=9.9, p<.001) in their average depression score. About half of the athletes (53.3%) who currently wanted or needed psychotherapeutic support for personal or mental health problems were classified as depressed. No differences were observed regarding eating disorders.

About a third of the athletes thought that ***harassment/abuse*** occurs in their sport ‘likely’ (n=83, 23.9%) or ‘very likely’ (n=42, 12.1%). Seventy-five athletes (21.6%) were ‘unsure’, 43 (12.4%) regarded this as ‘unlikely’, and 105 athletes (30.2%) thought that ‘no’ harassment/abuse occurs in their sport. The answers differed significantly between genders (Chi^2^=17.9, p<.01), with males regarding it more likely than females. Almost 80% of the athletes (n=273, 79.6%) stated that they have never witnessed or experienced themselves any forms of harassment/abuse in their sport environment. Fifty-one athletes (14.9%) had experienced it themselves, and 31 (9.0%) had witnessed it in another athlete (percentages include 12 athletes who had both) without difference between genders. Athletes with different experiences of harassment/abuse differed significantly (F=13.4, p<.001) on how likely they think it occurs in their sport. Athletes who had not witnessed nor experienced harassment/abuse regarded it as less likely than athletes who had witnessed it in another athlete or experienced it themselves, and those who had both experienced and witnessed it regarded it as most likely. Athletes with different experiences of harassment/abuse differed significantly in their average score of depression (F=4.6, p<.01, see Figure 1) and eating disorder screening tools (F=5.0, p<.01, see Figure 1) as well as on the subjective need for psychotherapeutic support (Chi^2^=15.4, p<.05). Athletes who experienced harassment/abuse themselves *and* witnessed it in another athlete had average lower values of depression and eating disorders than those who exclusively experienced it themselves (see Figure 1).

**Figure 1:** Average score on the depression (CESD-R-10, dark blue) and the eating disorder (BEDA-Q, light blue) scale in athletes with different experiences of harassment/abuse.

From a list of 13 *situations of harassment/abuse in the sports environment* (see Table 1) ‘unwanted comments about body or appearance’ was the most frequently (40.2%) and ‘rewards in sport for sexual favours’ (2.5%) and ‘rape’ (reported as ‘others’ n=1, 0.3%) the least frequently witnessed or experienced situation. In general, situations were more frequently witnessed in another athlete than experienced themselves, except for ‘inappropriate training’, ‘unsafe training conditions’, and ‘forced dieting, limited or no access to needed water or food’. About 10% of the athletes had experience these situations, which were reported to have been perpetrated most often by the coach. On average, one in twenty athletes (5.0%) had experienced themselves ‘unwanted and unnecessary touching of the body’ mostly by the coach or another athlete. Three athletes (0.8%) had experienced ‘rewards in sport for sexual favours’ themselves, and a further six (1.6%) had witnessed this in another athletes.

Significant more female than male athletes reported to have experienced harassment/abuse themselves with regard to ‘unwanted comments about body or appearance’ (38.5% vs 15.1%, Chi^2^=19.1, p<.001), ‘forced dieting, limited or no access to needed water or food’ (11.8% vs 1.5%, Chi^2^=12.3, p<.001), ‘bullying / cyber bullying’ (16.7% vs 5.3%, Chi^2^=9.4, p<.01), ‘unwanted and unnecessary touching of the body’ (7.3% vs 2.2%, Chi^2^=4.2, p<.01). About half of the athletes (n=181, 53.9%) reported they would talk to friends or family if they saw or experienced a situation like the ones listed in Table 1, but less than one in five (n=61, 18.8%) would talk to an official contact to seek help. About one in eight athletes (n=43, 12.8%) would not talk to anybody, and 68 (20.2%) were not certain whether or not to share the experience. The answers differed significantly between genders (F=22.7, p<.001). More than twice as many males than females (19.7% vs 8.7%) would not talk about it, and more females than males would talk to their friends and family (57.0%vs 33.9%).

### Specific analysis combining gender and discipline

were conducted for SW, WP, DV/HD (DV and HD combined) and female AS (see Table 2), since just three female and five male OWS and two male AS responded the survey.

**Table 2.** Characteristics of all elite aquatic athletes, and of male and female athletes from different selected disciplines (i.e. results from open water swimmers and male artistic swimmers not presented separately due to small numbers but included in total) and number (%) of athletes who experienced different types of harassment/abuse themselves with regard to gender and discipline.

|  | **Swimming** |  | **Water Polo** |  | **Diving / High Diving** |  | **Artistic Swimming** | **Total** |
| --- | --- | --- | --- | --- | --- | --- | --- | --- |
| Gender | female | male | female | male | female | male | female | both |
| Number of responding athletes | 65 | 67 | 48 | 34 | 29 | 34 | 77 | 375 |
| **Age** |  |  |  |  |  |  |  |  |
| Mean (sd) | 21.4 (4.9) | 22.3 (5.7) | 23.1 (3.9) | 24.8 (4.1) | 23.3 (5.9) | 26.7 (7.4) | 19.6 (3.8) | 22.4 (5.4) |
| **Depression (CES-D-10)** |  |  |  |  |  |  |  |  |
| Athletes with depression | 12 (27.3%) | 15 (31.9%) | 9 (27.3%) | 3 (12.5%) | 6 (23.1%) | 4 (15.4%) | 15 (28.3%) | 70 (26.1%) |
| **Eating disorder (BEDA-Q)** |  |  |  |  |  |  |  |  |
| Athletes with eating disorder | 16 (28.1%) | 14 (26.4%) | 16 (38.1%) | 8 (27.6%) | 11 (42.3%) | 8 (27.6%) | 33 (54.1%) | 111 (35.2%) |
| Lose weight now | 15 (24.2%) | 9 (14.8%) | 9 (19.6%) | 6 (18.8%) | 12 (41.4%) | 4 (12.5%) | 33 (44.0%) | 90 (25.3%) |
| **Need for psychotherapy** |  |  |  |  |  |  |  |  |
| No expressed need | 28 (45.2%) | 33 (55.0%) | 20 (42.6%) | 24 (72.7%) | 17 (58.6%) | 22 (71.0%) | 46 (61.3%) | 198 (55.6%) |
| Currently receiving | 16 (25.8%) | 14 (23.3%) | 4 (8.5%) | 5 (15.2%) | 5 (17.2%) | 3 (9.7%) | 7 (9.3%) | 57 (16.0%) |
| Previously received | 18 (29.0%) | 13 (21.7%) | 23 (48.9%) | 4 (12.1%) | 7 (24.1%) | 6 (19.4%) | 22 (29.3%) | 101 (28.4%) |
| **Harassment /abuse** |  |  |  |  |  |  |  |  |
| Likely in the athlete's sport<sup>(a)</sup> | 3.0 (1.4) | 3.2 (1.4) | 3.0 (1.4) | 3.3 (1.3) | 2.2 (1.4) | 2.4 (1.3) | 2.2 (1.2) | 2.8 (1.4) |
| Not experienced nor witnessed | 47 (75.8%) | 51 (87.9%) | 35 (77.8%) | 18 (58.1%) | 22 (84.6%) | 27 (87.1%) | 57 (81.4%) | 271 (79.5%) |
| Experienced themselves | 9 (14.5%) | 4 (6.9%) | 3 (6.7%) | 5 (16.1%) | 4 (15.4%) | 2 (6.5%) | 9 (12.9%) | 39 (11.4%) |
| Witnessed in another athlete | 3 (4.8%) | 2 (3.4%) | 5 (11.1%) | 5 (16.1%) | 0 | 1 (3.2%) | 2 (2.9%) | 19 (5.6%) |
| Experienced and witnessed | 3 (4.8%) | 1 (1.7%) | 2 (4.4%) | 3 (9.7%) | 0 | 1 (3.2%) | 2 (2.9%) |  |
| **Seek assistance** |  |  |  |  |  |  |  |  |
| No | 6 (10.0%) | 15 (25.9%) | 2 (4.7%) | 5 (17.2%) | 2 (8.0%) | 2 (6.7%) | 7 (9.9%) | 43 (12.9%) |
| Maybe | 10 (16.7%) | 16 (27.6%) | 8 (18.6%) | 7 (24.1%) | 5 (20.0%) | 8 (26.7%) | 13 (18.3%) | 68 (20.4%) |
| Friends or family | 35 (58.3%) | 11 (19.0%) | 25 (58.1%) | 14 (48.3%) | 12 (48.0%) | 13 (43.3%) | 41 (57.7%) | 161 (48.2%) |
| Official contact to seek help | 5 (8.3%) | 12 (20.7%) | 7 (16.3%) | 3 (10.3%) | 3 (12.0%) | 6 (20.0%) | 5 (7.0%) | 43 (12.9%) |
| Friends/family and official contact | 4 (6.7%) | 4 (6.9%) | 1 (2.3%) | 0 | 3 (12.0%) | 1 (3.3%) | 5 (7.0%) | 19 (5.7%) |
| **Athletes who experienced the specified type of harassment/abuse themselves** |  |  |  |  |  |  |  |  |
| Unwanted comments about body or appearance | 13 (20.6%) | 8 (11.9%) | 7 (24.9%) | 4 (12.1%) | 8 (32.0%) | 5 (15.2%) | 39 (55.7%) | 91 (25.2%) of 361 |
| Exclusion, isolation, or intentional denial of attention and/or support | 13 (21.0%) | 8 (12.1%) | 4 (8.5%) | 3 (9.1%) | 1 (3.8%) | 2 (6.1%) | 11 (16.4%) | 44 (12.4%) of 356 |
| Bullying / cyber bullying | 10 (16.1%) | 4 (6.0%) | 4 (8.7%) | 1 (3.0%) | 3 (11.5%) | 1 (3.0%) | 14 (20.3%) | 39 (10.8%) of 359 |
| Comments, jokes, messages, or gestures of a sexual nature | 9 (14.8%) | 5 (7.6%) | 4 (8.5%) | 3 (9.1%) | 2 (7.7%) | 1 (3.0%) | 4 (5.6%) | 30 (8.3%) of 361 |
| Negative comments on race, gender, religion, or country of origin | 3 (4.8%) | 2 (3.0%) | 3 (6.4%) | 2 (6.3%) | 2 (7.7%) | 0 | 7 (9.9%) | 22 (6.1%) of 361 |
| Inappropriate training, e.g. forced physical exertion beyond capabilities | 9 (14.8%) | 6 (9.1%) | 3 (6.5%) | 4 (12.1%) | 0 | 0 | 13 (18.8%) | 39 (10.9%) of 357 |
| Intentional hitting, beating, or kicking (not fouls or rough | 2 (3.2%) | 5 (7.5%) | 3 (6.4%) | 3 (9.1%) | 0 | 0 | 4 (5.9%) | 18 (5.0%) of 360 |
| play) |  |  |  |  |  |  |  |  |
| Forced dieting, limited or no access to needed water or food | 9 (14.3%) | 0 | 2 (4.3%) | 1 (3.0%) | 1 (3.8%) | 0 | 12 (17.1%) | 27 (7.5%) of 361 |
| Unsafe training conditions | 7 (11.3%) | 4 (6.2%) | 4 (8.7%) | 1 (3.0%) | 5 (19.2%) | 2 (6.1%) | 5 (7.1%) | 30 (8.4%) of 358 |
| Unwanted and unnecessary touching of the body | 8 (12.9%) | 3 (4.5%) | 4 (8.5%) | 0 | 0 | 0 | 3 (4.3%) | 18 (5.0%) of 360 |
| Unwanted touching of breast, buttock, genitals or attempt to kiss | 5 (7.9%) | 2 (3.0%) | 1 (2.1%) | 0 | 0 | 0 | 2 (2.9%) | 11 (3.0%) of 361 |
| Forced use of supplements / medication | 3 (4.8%) | 1 (1.5%) | 1 (2.1%) | 1 (3.0%) | 0 | 0 | 0 | 7 (1.9%) of 362 |
| Rewards in sport for sexual favours | 2 (3.2%) | 1 (1.5%) | 0 | 0 | 0 | 0 | 0 | 3 (0.8%) of 362 |
| Other, please specify | 1 (1.9%) | 0 | 0 | 0 | 0 | 0 | 0 | 2 (0.6%) of 327 |

Numbers do not add up to responding athletes due to missing values in gender (n=2), discipline (n=12) and the respective question. (a) no =1; not likely =2; not sure =3; likely =4; very likely =5.

*Gender differences within a discipline*, did not reach statistical significance due to the small subgroups, except for ‘trying to lose weight now’ in DV/HD (Chi^2^=6,6, p<.05), need for psychotherapy in WP (Chi^2^=11.8, p<.01), ‘talk about experience/witnessed harassment/abuse’ in SW (Chi^2^=20.6, p<.001), and ‘experienced themselves forced dieting, limited or no access to needed water or food’ in SW (Chi^2^=10.1, p≤.001). However, in several variables, a substantially higher proportion of female than male athletes was observed, e.g. athletes with an eating disorder in WP and DV/HD, ‘try to lose weight now’ in SW, experienced harassment/abuse themselves in SW and HD (see Table 2). Further, it is interesting to note that a similar proportion of female and male SW reported a current need for psychotherapy, and that in WP more males than females had experienced harassment/abuse themselves. The latter concerns specifically ‘inappropriate training’ and ‘intentional hitting, beating, or kicking (not fouls or rough play)’.

Comparing *female athletes* competing in AS, SW, DV/HD and WP, significant differences were observed with regard to eating disorders (Chi^2^=8.4, p<.05), ‘try to lose weight now’ (Chi^2^=11.0, p<.05) and the need for psychotherapeutic support (Chi^2^=15.6, p<.05). Also, age (F=8.1, p<.001) and the perception of the occurrence of harassment/abuse in their sport (F=5.8, p<.01) differed significantly between the four disciplines (see Table 2). Significant differences between disciplines were observed for female athletes who experiences themselves ‘unwanted comments about body or appearance’ (Chi^2^=27.8, p<.001) ranging from AS (n=39, 55.7%) to WP (n=7; 14.9%), and ‘inappropriate training’ (Chi^2^=8.1, p<.05) ranging from AS (n=13, 18.8%) to DV/HD (n=0). ‘Forced dieting, limited or no access to needed water or food’ ranged from AS (n=13, 17.1%) to DV/HD (n=1, 3.8%) but the difference did not reach statistical significance (Chi^2^=6.5, p<.09) (see Table 2). In five of the 13 situations of harassment/abuse, female AS had the highest percentage of affected athletes, and in four female SW.

Comparing *male athletes* between three disciplines, age (F=6.6, p<.01), the perception of the occurrence of harassment/abuse in their sport (F=4.6, p<.05) and the proportion of athletes who had witnessed or experienced harassment/abuse (Chi^2^=13.5, p<.05) differed significantly (see Table 2). Further differences, such as more SW than WP or DV/HD were classified as depressed (31.9% vs 12.5% resp 15.4%) and needed psychotherapeutic support currently (23.3% vs 15.2% resp. 9.7%), were not statistically significant. Also differences in the 13 situations of harassment/abuse did not reach significance between disciplines in male athletes, most probably because of small numbers (see Table 2).

## DISCUSSION

This study is the first to evaluate mental health and harassment/abuse in elite aquatic athletes. About a quarter of athletes were classified as **depressed**, which is consistent with previous publications.^20-22^ In the general population and other studies on athletes,^21,23^ more female than male athletes reported symptoms of depression. This also applies to the present study, except for swimmers. The prevalence of depression in male swimmers was about twice as high as in male athletes of the other aquatic disciplines. Our study demonstrated a significant correlation between the depression score and the athletes *i)* symptoms/health complaints and *ii)* their effect on performance in the four weeks prior to the WC. The intersection between injury and mental health is multi-faceted as elite sport can have stressors that increase the risk of both mental health disorders and injury. In addition, injury may trigger mental health symptoms while conversely, mental health disorders may prolong recovery from injury.^4^

More than a third of the athletes were screened as having an **eating disorder**, with a higher proportion of female (40%) than male athletes (27%), and highest in AS (54.1%). This finding is not surprising as AS is an aesthetic judged sport, where a quintessential lean body composition is implicitly valued in the culture of the sport.^24^ Interestingly, in SW, there were equal proportions of female and males (28.1% vs 26.4%) with eating disorders which could be related to the gender-neutral importance of body composition on performance (decreasing drag), rather than the more common culturally-driven body image reasons that more often influence female than male athletes. Overall, a quarter of the athletes stated that they were trying to lose weight during the WC, however this applied to more than 40% of females in the two judged disciplines (AS, DV/HD). This finding is disturbing with almost half of the competing athletes in the highest competition level in the world engaging in weight loss behaviours that could affect their nutritional fuelling of performance. It also has significant medical implications for team physicians with the associated risks of weight loss during highly competitive sport. Interestingly, in WP, the male and female reporting of current weight loss behaviours was similar (about 20%), which is high for males and low for females. The estimated prevalence of eating disorders and/or disordered eating among athletes in general, ranges from 0% to 19% in males and from 6% to 45% in females.^25^

In the present study, 45% of participating athletes wanted or needed **psychotherapeutic support** for personal or mental health problems either previously (29%) or currently (16%). Similar results were reported in a survey on elite female football players (24% previous; 16% currently).^18^ Consistent with the high prevalence of depression in male swimmers, approximately 25% of both female and male swimmers were *currently* wanting or needing professional mental health support. These findings underscore the need for team physicians of elite athletes to have the core clinical competency to diagnose and treat mental health disorders^26^, and for organizing committees and International Federations to incorporate mental health professionals into the medical care services for large multi-sport events.^27^

Over one-third (36%) of athletes participating in the present study stated that **harassment/abuse** was either ‘very likely’ or ‘likely’ to occur in their sport. Similar percentages were found in youth athletes participating in the Youth Olympic Games 2018.^10^ In the present study 15% of the athletes reported to have experienced harassment/abuse in their sports environment themselves. For specific situations, the frequency ranged from ‘unwanted comments about body or appearance’ (40.2%) to ‘rewards in sport for sexual favours’ (2.5%) and ‘rape’ (0.3%). A study of German and Belgian-Dutch elite athletes found a prevalence of psychological (72%), physical (25%) and sexual harassment/abuse (31%) with significant overlap of types.^28^ A cross-sectional study of 198 elite Swedish athletics athletes reported a sexual abuse prevalence of 11% (females 16% and males 4%) and physical abuse of 18% (females 14% and males 23%).^29^ A survey of 1001 elite Canadian athletes from 64 sports reported a prevalence of 59% for emotional harm, 67% for neglect, 20% for sexual harm and 12% for physical harm.^6^ In the present study, athletes who have experienced harassment or abuse in their sports environment themselves had higher average scores of the depression and of eating disorders, and more of them needed psychotherapeutic support than those who have not witnessed nor experienced it. While our findings are consistent with reports in the literature, they remain alarming. Validated prevention strategies to decrease harassment/abuse in the aquatic sports are urgently required.

In this survey, only 19% stated that they would **officially report harassment/abuse**, while 13% would say nothing, and 20% were unsure what to do. These numbers are consistent with the Canadian study who found that those athletes who had experienced harassment/abuse, 56% did not report their experience, and 84% did not submit a formal complaint.^6^ These findings demonstrate the need *i)* to encourage athletes to report their abuse, *ii)* to develop safe reporting mechanisms, and *iii)* to ensure experienced athlete support systems are accessible.

There are **limitations** to this study inherent in the retrospective design including selection and recall bias. In addition, the response rate was low which could result in an over- or underestimation of the prevalence.^30^ However, we believe that a selection bias is unlikely, since responses to the questions on health complaints in the 4 weeks prior to the WC were similar to the athlete surveys in FINA WC 2013.^11-13^

## CONCLUSIONS

This novel study has uncovered the stark realities that lie beneath the surface of the water for elite aquatic athletes. Attention is needed to improve understanding through targeted research regarding differences between genders and disciplines, as well as to develop educational initiatives to decrease the stigma of mental health in sport. Mechanisms are required to improve athlete help-seeking, to facilitate reporting of harassment/abuse, and to enhance athlete support systems. Prevention strategies should be implemented and evaluated to address both athlete mental health challenges and harassment/abuse in the aquatic sports. The results of this study should serve as an alarm to engage and empower athletes and their entourage to work together with FINA to address athlete mental well-being and Safe Sport in the daily training environments of the swimming pools around the world.

> *"There are times where I feel absolutely worthless, where I completely shut down but have this bubbling anger that is through the roof. If I’m being honest, more than once I’ve just screamed out loud, “I wish I wasn’t me!*… *The more you understand that it’s ok, to not be ok – it’s so powerful”* Michael Phelps (2020)^31^

## PRACTICAL IMPLICATIONS

- Sport organizations should develop strategies to decrease stigma related to athlete mental health problems.
- Team physicians should be educated to have the clinical competency to screen, diagnose and treat mental health problems in elite aquatic athletes regarding differences between genders and disciplines.
- Prevention strategies should be implemented to decrease the burden of mental health issues and harassment/abuse in sport.
- Reporting mechanisms and athlete support systems should be made available and accessible for athletes experiencing harassment/abuse in sport.

## Data Availability

The data is currently not publically available.

## Acknowledgement

The authors greatly appreciate the cooperation of the athletes attending the FINA World Aquatic Championships who volunteered their time to provide data for this project, and to FINA for supporting the concept and logistics of the study.

## Abbreviations

AS: Artistic swimming
BEDA-Q: Brief Eating Disorders in Athletes Questionnaire
CES-D-10: Center for Epidemiological Studies Depression Scale revised
DV: Diving
FINA: Federation Internationale de Natation
HD: High Diving
IOC: International Olympic Committee
OWS: Open water swimming
SW: Swimming
WC: World Championships
WP: Water polo

## REFERENCES

1. Scutti S. “Michael Phelps: I am extremely thankful that I did not take my life” CNN 20 Jan 2018. Available: https://www.cnn.com/2018/01/19/health/michael-phelps-depression/index.html [Accessed 22 Sep 2020]

2. Drehs W. Michael Phelps’ final turn. 2016. EPSN the Magazine. Available: https://www.espn.com/espn/feature/story/_/id/16425548/michael-phelps-prepares-life-2016-rio-olympics [Accessed 22 Sep 2020]

3. Gouttebarge V, Castaldelli-Maia JM, Gorczynski P, et al. Occurrence of mental health symptoms and disorders in current and former elite athletes: a systematic review and meta-analysis. Br J Sports Med. 2019;53:700–6. http://dx.doi.org.libaccess.lib.mcmaster.ca/10.1136/bjsports-2019-100671

4. Reardon C, Hainline B, Aron CM, et al. Mental health in elite athletes: International Olympic Committee consensus statement (2019). Br J Sports Med 2019:53(11):667–99. http://dx.doi.org/10.1136/bjsports-2019-100715

5. Timpka T, Spreco A, Dahlstrom et al. Suicidal thoughts (ideation) among elite athletics (track and field) athletes: cross-sectional study of associations with sexual and physical abuse victimization, aspects of sports participation, and psychological and behavioural resourcefulness. Br J Sports Med 2020 https://doi:10.1136/bjsports-2019-101386

6. Kerr G, Willson E, Stirling A. Prevalence of maltreatment among current and former national team athletes. University of Toronto in partnership with Athletes CAN 20 April 2019. Available: http://athletescan.com/sites/default/files/images/prevalence_of_maltreatment_reporteng.pdf [Accessed 22 Sep 2020]

7. International Olympic Committee. Consensus Statement on “Sexual harassment and abuse in sport”. February 2007. Available: https://www.olympic.org/news/ioc-adopts-consensus-statement-on-sexual-harassment-and-abuse-in-sport [Accessed 22 Sep 2020]

8. International Olympic Committee Safeguarding Framework, Rio de Janeiro Olympic Games 2016. Available: https://cdn.dosb.de/alter_Datenbestand/Bilder_allgemein/Veranstaltungen/Rio2016/IOC_Framework_for_safeguarding_athletes.pdf [Accessed 22 Sep 2020]

9. Mountjoy M, Brackenridge C, Arrington M, et al. The IOC Consensus Statement: Harassment and abuse (non-accidental violence) in sport. Br J Sports Med 2016; (17):1019–29. https://doi.org/10.1136/bjsports-2016-096121

10. Mountjoy M, Vertommen T, Burrows K, et al., #SafeSport: Safeguarding initiatives at the Youth Olympic Games 2018. Br J Sports Med 2020;54:176–82. https://doi.org/10.1136/bjsports-2019-101461

11. Mountjoy M, Junge A, Benjamen S, et al. Competing with injuries: injuries prior to and during the 15th FINA World Championships 2013 (aquatics). Br J Sports Med. 2015;49(1):37–43. http://dx.doi.org/10.1136/bjsports-2014-093991

12. Prien A, Mountjoy M, Miller J, et al. Injury and illness in aquatic sport –how is the risk? A comparison of results from three FINA World Championships. Br J Sports Med 2017;51(4):277–82. http://dx.doi.org/10.1136/bjsports-2016-096075

13. Mountjoy M, Junge A, Slysz J, et al. An uneven playing field: Athlete injury, illness, load, and daily training environment in the year prior to the FINA World Championships, 2017. [published online ahead of print, 2019 Dec 12]. Clin J Sport Med 2019;. https://doi:10.1097/JSM.0000000000000814

14. Clarsen B, Rønsen O, Myklebust G, et al. The Oslo Sports Trauma Research Center questionnaire on health problems: a new approach to prospective monitoring of illness and injury in elite athletes. Br J Sports Med 2014;48:754–60. http://dx.doi.org/10.1136/bjsports-2012-092087

15. Björgvinsson T, Kertz SJ, Bigda-Peyton JS, et al. Psychometric properties of the CES-D-10 in a psychiatric sample. Assessment 2013, 20, 429–36. https://doi.org/10.1177/1073191113481998

16. Zhang W, O’Brien N, Forrest JI, et al. Validating a shortened depression scale (10 item CES-D) among HIV-positive people in British Columbia, Canada. PLoS One. 2012;7(7):e40793. https://doi.org/10.1371/journal.pone.0040793

17. Martinsen M, Holme I, Pensgaard AM, et al. The development of the Brief Eating Disorder in Athletes Questionnaire (BEDA-Q). Med Sci Sport Exer 2014;46(8):1666–75. https://doi.org/10.1249/mss.0000000000000276

18. Junge A, Prinz B. Depression and anxiety symptoms in 17 teams of female football players including 10 German first league teams. Br J Sports Med 2019;53:471–7. http://dx.doi.org/10.1136/bjsports-2017-098033

19. Timpka T, Janson S, Jacobsson J. et al.. Protocol design for large-scale cross-sectional studies of sexual abuse and associated factors in individual sports: Feasibility study in Swedish athletics. J Sport Sci Med 2015;14:179-87. PMID: 25729306; PMCID: PMC4306771

20. Armstrong S, Oomen-Early J. Social connectedness, self-esteem, and depression symptomatology among collegiate athletes versus non-athletes. J Am Coll Health 2009;57:521–6. https://doi.org/10.3200/JACH.57.5.521-526

21. Gulliver A, Griffiths KM, Mackinnon A, et al. The mental health of Australian elite athletes. J Sci Med Sport 2015;18:255–61. https://doi.org/10.1016/j.jsams.2014.04.006

22. Yang J, Peek-Asa C, Corlette JD, et al. Prevalence of and risk factors associated with symptoms of depression in Competitive Collegiate Student Athletes. Clin J Sport Med 2007;17:481–7. https://doi:10.1097/JSM.0b013e31815aed6b

23. Junge A, Feddermann-Demont N. Prevalence of depression and anxiety in top-level male and female football players. BMJ Open Sport Exerc Med 2016;2:e000087. http://dx.doi.org/10.1136/bmjsem-2015-000087

24. Ferrand C, Magnan C, Rouveix M, et al. Disordered eating, perfectionism and body-esteem of elite synchronized swimmers. Eur J Sport Sci 2007: 223–30. http://doi:10.1080/17461390701722168

25. Bratland-Sanda S, Sundgot-Borgen J. Eating disorders in athletes: overview of prevalence, risk factors and recommendations for prevention and treatment. Eur J Sport Sci 2013;13:499–508. https://doi.org/10.1080/17461391.2012.740504

26. Gouttebarge V, Bindra A, Blauwet C, et al. International Olympic Committee (IOC) Sport Mental Health Assessment Tool 1 (SMHAT-1) and Sport Mental Health Recognition Tool 1 (SMHRT-1): towards better support of athletes’ mental health. Br J Sports Med. 2020:bjsports-2020-102411. http://doi:10.1136/bjsports-2020-10241

27. Mountjoy M, Moran J, Amed H, et al. Athlete health and safety at large sport events: the development of consensus-driven guidelines. Br J Sports Med 2020. [in publication]

28. Ohlert J, Vertommen T, Rulofs B, et al., Elite athletes’ experiences of interpersonal violence in organised sport in Germany, the Netherlands, and Belgium. Europ J Sport Sci 2020; 1–10. [published online ahead of print, 2020 Jul 7]. https://doi:10.1080/17461391.2020.1781266

29. Timpka T, Janson S, Jacobsson J et al. Lifetime sexual and physical abuse among elite athletics athletes: A cross-sectional study of prevalence and correlates with athletics injury. Br J Sports Med 2014;48:667. http://dx.doi.org/10.1136/bjsports-2014-093494.285

30. Kawakami N, Yasuma N, Watanabe K, et al. Association of response rate and prevalence estimates of common mental disorders across 129 areas in a nationally representative survey of adults in Japan. Soc Psychiatry Psychiatr Epidemiol. 2020: https://doi.org/10.1007/s00127-020-01847-3

31. Phelps M. as told to Drehs W. „Michael Phelps:, Thi is the most overwhelmed I’ve ever felt’. ESPN 18 May 2020. Available: https://www.espn.com/olympics/story/_/id/29186389/michael-phelps-most-overwhelmed-ever-felt [Accessed 22 Sep 2020]

